# Changing patterns of sickness absence among healthcare workers in England during the COVID-19 pandemic

**DOI:** 10.1101/2021.04.08.21255128

**Authors:** Rhiannon Edge, Diana A van der Plaat, Vaughan Parsons, David Coggon, Martie van Tongeren, Rupert Muiry, Ira Madan, Paul Cullinan

## Abstract

**Objective:** To explore impacts of the COVID-19 pandemic on patterns of sickness absence among staff employed by the National Health Service (NHS) in England.

**Methods:** We analysed prospectively collected, pseudonymised data on 959,356 employees who were continuously employed by NHS trusts during 1 January 2019 to 31 July 2020, comparing the frequency of new sickness absence in 2020 with that at corresponding times in 2019.

**Results:** After exclusion of episodes directly related to COVID-19, the overall incidence of sickness absence during the initial 10 weeks of the pandemic (March-May 2020) was more than 20% lower than in corresponding weeks of 2019, but trends for specific categories of illness varied. Marked increases were observed for asthma (122%), infectious diseases (283%) and mental illness (42.3%), while reductions were apparent for gastrointestinal problems (48.4%), genitourinary/gynaecological disorders (33.8%), eye problems (42.7%), injury and fracture (27.7%), back problems (19.6%), other musculoskeletal disorders (29.3%), disorders of ear, nose and throat (32.7%), cough/flu (24.5%) and cancer (24.1%). A doubling of new absences for pregnancy-related disorders during 18 May to 19 July of 2020 was limited to women with earlier COVID-19 sickness absence.

**Conclusions:** Various factors will have contributed to the large and divergent changes that were observed. The findings add to concerns regarding delays in diagnosis and treatment of cancers, and support a need to plan for a large backlog of treatment for many other diseases. Further research should explore the rise in absence for pregnancy-related disorders among women with earlier COVID-19 sickness absence.

**1. What is already known about this subject?**
Historically, rates of sickness absence among the NHS workforce in England have been relatively high but stable. Reports of a marked increase during the first wave of the COVID-19 pandemic have not distinguished between different categories of underlying illness.
**2. What are the new findings?**
During the first wave of COVID-19, incidence of sickness-absence changed markedly when compared to the previous year, with major increases for some categories of illness, and large declines for many others, including cancer.
**3. How might this impact on policy or clinical practice in the foreseeable future?**
The findings support a need to plan for effects from delayed diagnosis and treatment of cancer, and to manage a large backlog of treatment for many other diseases.

## Introduction

In addition to its impacts on productivity, sickness absence from work is important for the light that it sheds on patterns of disease and illness-related behaviours in working populations. During the COVID-19 pandemic, healthcare workers have faced a serious threat to their personal safety, in combination with new and heightened occupational demands from an uncertain and rapidly evolving crisis. Rates of infection by SARS-CoV-2 have been higher in healthcare workers than in most other occupations.(1-3) Workload has been increased not only by the exceptional number of patients requiring treatment for the disease, but also by a need to cover for colleagues, who were themselves infected, isolating because of possible close contact with infection, or shielding from exposure to infection because of personal vulnerability. A challenge of this sort could be expected to affect sickness absence in diverse ways. In addition, the pandemic has had wider effects on people’s activities and access to health services which could further alter patterns of sickness absence (e.g. through postponement of less urgent clinical investigation and treatment).

Before the pandemic, staff employed by the National Health Service (NHS) in England had persistently high, but stable, overall rates of sickness absence.(4, 5) Preliminary analyses have indicated a sudden rise during the early phase of the epidemic nationally (March - April 2020) with notable variability across geographical areas and between staff groups.(6-8) Much of this increase will have been driven by absence because of COVID-19, but to gain a better understanding, it is necessary to break down trends according to different medical reasons for absence.

In this paper, we report an analysis of data on NHS employees in England, to explore the nature and extent of changes in sickness absence for categories of health problem other than COVID-19 during the first wave of the pandemic. We also examine whether absence for COVID-19 infection during the early weeks of the epidemic was associated with altered patterns of sickness absence for other types of illness in the longer term.

## Methods

We analysed pseudonymised data that had been abstracted on our behalf from the National Health Service (NHS) Electronic Staff Record (ESR). The ESR is a centralised database to which all NHS trusts in England contribute monthly-updated personnel records, using a standardised coding system. The information to which we were given access related to all staff who had been continuously employed by NHS trusts in England between 1 January 2019 and 31 July 2020. It included demographic and occupational characteristics for each individual, together with data on all absences from work during that period (other than for annual leave), detailing the dates that each episode started and finished, and the reason for absence. A more extensive description of the source material and its preliminary processing has been presented in an earlier report.(9)

For the current analysis we focused on numbers of new episodes of sickness absence across the whole of the study sample, classified according to the week of the year in which they started, their duration (≤7 days or >7 days) and the reason for absence. Reason for sickness absence was classified to 22 diagnostic categories, as in the source ESR database. In addition, since 17 March 2020, trusts had been given the option to record whether an absence was related to COVID-19, using a code that was entered as an adjunct to other diagnostic categories.

Statistical analysis was carried out using R Statistical Software (version 4.0.4).(10) We calculated percentage changes (with 95% confidence intervals (CIs)) from 2019 to 2020 in the numbers of new episodes of sickness absence during corresponding weeks of the year. The main periods that we examined (Table 1) were chosen to cover the time immediately before the first wave of the COVID-19 epidemic in England took off (weeks 2-10), the time when it was at its height (weeks 11-20), and then a time when it was subsiding (weeks 21-29). Year-on-year changes were assessed for all sickness absence, and for sickness absence in which there was no record of COVID-19 as a related reason.

**Table 1.** Main periods studied.

| **Weeks** | **2019** | **2020** |
| --- | --- | --- |
| 2-10 | 7 January to 10 March | 6 January to 8 March |
| 11-18 |  | 9 March to 3 May |
| 11-20 | 11 March to 19 May | 9 March to 17 May |
| 21-29 | 20 May to 21 July | 18 May to 19 July |

To explore whether infection by COVID-19 was associated with altered patterns of sickness absence in the longer term, we also analysed year-on-year changes for specific categories of absence during weeks 21-29, according to whether individuals had taken COVID-19 sickness absence during weeks 11-18 of 2020. For this purpose, COVID-19 sickness absence was defined as in earlier reports (9) – i.e. sickness absence in any of five diagnostic categories (cough/flu, chest/respiratory, infectious diseases, other, unknown) with COVID-19 recorded as a related reason.

Ethical approval for the study was provided by the NHS Health Research Authority (reference 20/SC/0282).

## Results

After exclusion of 21,775 individuals who were absent continuously throughout the study period (e.g. for study leave), analysis was based on 959,356 employees. Most (89%) were aged between 25 and 60 years, and 77% were female.

Table 2 shows the numbers of new episodes of sickness absence within the study sample during corresponding nine- or ten-week periods in 2019 and 2020, and the percentage change from the first to the second year for each period. Across the three periods in 2020, COVID-19 was recorded as a related reason for absence in a total of 101,585 new episodes, of which 100,833 (99%) met our specified criteria for COVID-19 sickness absence. Most (87%) began during weeks 11 to 20, and only 414 (0.4%) started earlier in the year.

**Table 2.** Changes in numbers of new episodes of sickness absence from 2019 to 2020 by period of year and category of sickness absence.

| Category of sickness absence | Weeks 2-10 |  |  | Weeks 11-20 |  |  | Weeks 21-29 |  |  |
| --- | --- | --- | --- | --- | --- | --- | --- | --- | --- |
|  | Number in 2019 | Number in 2020 | <sup>a</sup>Percentage change (95%CI) | Number in 2019 | Number in 2020 | <sup>a</sup>Percentage change (95%CI) | Number in 2019 | Number in 2020 | <sup>a</sup>Percentage change (95%CI) |
| All categories | 278,006 | 275,134 | -1.0 (-1.6 to -0.5) | 250,011 | 284,935 | 14.0 (13.4 to 14.6) | 218,190 | 170,005 | -22.1 (-22.6 to -21.6) |
| COVID-19<sup>b</sup> |  | 414 |  |  | 88,695 |  |  | 12,476 |  |
| All categories except COVID-19 | 278,006 | 274,720 | -1.2 (-1.7 to -0.7) | 250,011 | 196,240 | -21.5 (-22.0 to -21.0) | 218,190 | 157,529 | -27.8 (-28.3 to -27.3) |
| Asthma | 1,057 | 1,113 | 5.3 (-3.2 to 14.5) | 959 | 2,130 | 122 (106 to 140) | 885 | 781 | -11.8 (-19.8 to -2.8) |
| Back problems | 9,409 | 10,130 | 7.7 (4.7 to 10.7) | 10,459 | 8,410 | -19.6 (-21.9 to -17.2) | 9,859 | 9,622 | -2.4 (-5.1 to 0.4) |
| Blood disorder | 526 | 577 | 9.7 (-2.5 to 23.5) | 646 | 476 | -26.3 (-34.5 to -17.1) | 640 | 427 | -33.3 (-41 to -24.6) |
| Cancer | 981 | 958 | -2.3 (-10.7 to 6.7) | 1,041 | 790 | -24.1 (-30.8 to -16.8) | 1,053 | 678 | -35.6 (-41.5 to -29.1) |
| Cardiac and circulatory | 2,315 | 2,528 | 9.2 (3.2 to 15.5) | 2,673 | 2,525 | -5.5 (-10.5 to -0.3) | 2,566 | 2,403 | -6.4 (-11.4 to -1.0) |
| Chest and respiratory | 12,787 | 11,725 | -8.3 (-10.6 to -6.0) | 9,143 | 12,083 | 32.2 (28.6 to 35.8) | 6,863 | 3,298 | -51.9 (-53.9 to -49.9) |
| Cough, flu | 82,504 | 72,641 | -12.0 (-12.8 to -11.1) | 45,138 | 34,070 | -24.5 (-25.6 to -23.5) | 28,110 | 6,403 | -77.2 (-77.8 to -76.6) |
| Dental and oral problems | 3,133 | 3,371 | 7.6 (2.5 to 13.0) | 3,633 | 2,391 | -34.2 (-37.5 to -30.7) | 3,418 | 2,929 | -14.3 (-18.4 to -10.0) |
| Ear, nose, throat | 9,773 | 10,831 | 10.8 (7.8 to 13.9) | 10,181 | 6,854 | -32.7 (-34.7 to -30.6) | 9,074 | 5,377 | -40.7 (-42.7 to -38.7) |
| Endocrine, glandular problems | 791 | 851 | 7.6 (-2.3 to 18.5) | 943 | 662 | -29.8 (-36.4 to -22.5) | 887 | 654 | -26.3 (-33.4 to -18.4) |
| Eye problems | 2,644 | 2,895 | 9.5 (3.9 to 15.4) | 3,108 | 1,782 | -42.7 (-45.9 to -39.2) | 2,878 | 2,001 | -30.5 (-34.3 to -26.4) |
| Gastrointestinal problems | 50,359 | 49,775 | -1.2 (-2.4 to 0.1) | 54,410 | 28,098 | -48.4 (-49.1 to -47.6) | 49,317 | 32,687 | -33.7 (-34.6 to -32.8) |
| Genitourinary, gynaecological | 7,405 | 8,072 | 9.0 (5.6 to 12.5) | 8,188 | 5,417 | -33.8 (-36.1 to -31.5) | 7,964 | 6,327 | -20.6 (-23.1 to -17.9) |
| Headache, migraine | 15,255 | 17,210 | 12.8 (10.4 to 15.3) | 17,239 | 16,358 | -5.1 (-7.1 to -3.1) | 17,362 | 19,053 | 9.7 (7.5 to 12.0) |
| Infectious diseases | 1,644 | 1,699 | 3.3 (-3.4 to 10.6) | 1,670 | 6,401 | 283 (263 to 305) | 1,690 | 1,454 | -14.0 (-19.8 to -7.7) |
| Injury, fracture | 5,797 | 6,214 | 7.2 (3.4 to 11.1) | 6,681 | 4,832 | -27.7 (-30.3 to -24.9) | 6,634 | 5,809 | -12.4 (-15.5 to -9.3) |
| Mental health | 15,888 | 20,470 | 28.8 (26.2 to 31.5) | 17,749 | 25,259 | 42.3 (39.6 to 45.1) | 17,974 | 20,792 | 15.7 (13.4 to 18.0) |
| Nervous system disorders | 971 | 1,089 | 12.2 (2.9 to 22.3) | 1,095 | 891 | -18.6 (-25.5 to -11.1) | 1,082 | 950 | -12.2 (-19.5 to -4.2) |
| Other musculoskeletal disorders | 12,630 | 13,478 | 6.7 (4.2 to 9.3) | 14,137 | 9,997 | -29.3 (-31.1 to -27.5) | 14,282 | 11,748 | -17.7 (-19.7 to -15.7) |
| Pregnancy-related disorders | 7,151 | 7,082 | -1.0 (-4.2 to 2.3) | 6,211 | 5,179 | -16.6 (-19.6 to -13.5) | 3,699 | 4,079 | 10.3 (5.5 to 15.3) |
| Skin disorders | 1,702 | 1,854 | 8.9 (2.0 to 16.3) | 2,007 | 1,892 | -5.7 (-11.5 to 0.4) | 2,248 | 2,066 | -8.1 (-13.4 to -2.4) |
| Other and unknown | 33,284 | 30,130 | -9.5 (-10.9 to -8.1) | 32,700 | 19,743 | -39.6 (-40.7 to -38.5) | 29,705 | 17,991 | -39.4 (-40.5 to -38.3) |

<sup>a</sup>Calculated as 100\*(Number in 2020 – Number in 2019)/Number in 2019
<sup>b</sup>Any sickness absence in which COVID-19 was recorded as a related reason for absence

For sickness absence that was not recorded as COVID-related, the overall number of new episodes during weeks 2-10 of 2020 was similar to that in the corresponding weeks of 2019 (274,720 vs. 278,006), although within that, there were increases for mental illness (by 28.8%) and headache/migraine (by 12.8%), offset by a reduction for cough/flu (by 12.0%). In contrast, much larger year-on-year changes were observed during weeks 11-20. The total number of new non-COVID absences fell by 21.5%, including reductions for gastrointestinal problems (by 48.4%), genitourinary/gynaecological disorders (by 33.8%), eye problems (by 42.7%), injury and fracture (by 27.7%), back problems (by 19.6%), other musculoskeletal disorders (by 29.3%), disorders of ear, nose and throat (by 32.7%), cough/flu (by 24.5%) and cancer by (24.1%). On the other hand, large increases were observed for infectious diseases (by 283%), asthma (by 122%), chest and respiratory disorders (by 32.2%) and mental illness (by 42.3%). In the third period (weeks 21-29), the overall year-on-year reduction in non-COVID absences was maintained (down by 27.8%), with changes for most specific diagnostic categories in the same direction as for weeks 11-20. Exceptions, however, were asthma, chest and respiratory disorders and infectious diseases, for all of which numbers were lower in 2020 than in 2019, and pregnancy-related disorders, for which there was a 10% increase.

To explore the timing of changes in more detail, Figure 1 shows the percentage change from 2019 to 2020 in new episodes of sickness absence by individual week of the year for selected diagnostic categories, and also weekly numbers of new episodes of COVID-19 sickness absence during 2020. The surges in absence for infectious diseases, asthma and chest and respiratory disorders all coincided closely with the emergence of absences for COVID-19, peaking 1 to 2 weeks earlier, while the increase in new absences for mental illness was less steep and peaked several weeks later.

**Figure 1.**
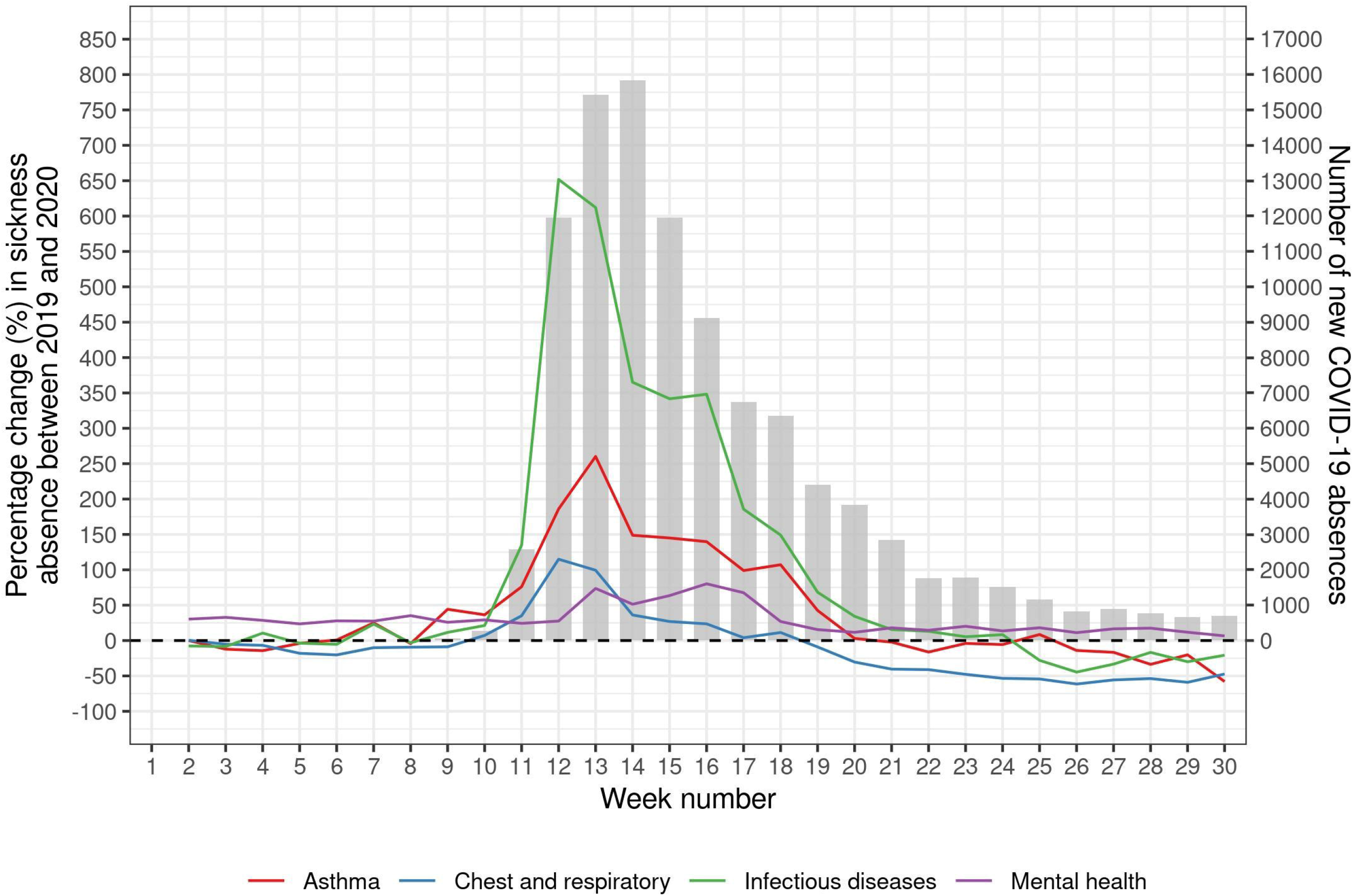
Percentage change from 2019 to 2020 in new episodes of sickness absence for selected causes and number of new absences for COVID-19 by week of year.

Table 3 breaks down the year-on-year changes in numbers of new absence episodes during weeks 11-29 according to whether they were of short (≤7 days) or longer duration. The increases for asthma, chest and respiratory disorders, infectious diseases and mental illness were all larger for longer-term than for short-duration episodes. There were also increases in long-duration absences for cough/flu and cardiac and circulatory disease, whereas short-duration absences for these diagnostic categories were less frequent in 2020 than in 2019. The reduction in absences for cancer was greater for short-duration episodes (48.1%), but was apparent also for episodes of longer duration (18.0%).

**Table 3.** Changes in numbers of new episodes of sickness absence during weeks 11-29 from 2019 to 2020 by category and duration of sickness absence.

| Category of sickness absence | Duration of absence ≤7 days |  |  | Duration of absence >7 days |  |  |
| --- | --- | --- | --- | --- | --- | --- |
|  | Number in 2019 | Number in 2020 | <sup>a</sup>Percentage change (95%CI) | Number in 2019 | Number in 2020 | <sup>a</sup>Percentage change (95%CI) |
| All categories | 379,770 | 300,545 | -20.9 (-21.2 to -20.5) | 88,431 | 154,395 | 74.6 (73.2 to 76.0) |
| All categories except COVID-19 | 379,770 | 250,449 | -34.1 (-34.4 to -33.7) | 88,431 | 103,320 | 16.8 (15.8 to 17.9) |
| Asthma | 1,471 | 1,647 | 12.0 (4.4 to 20.1) | 373 | 1,264 | 239 (202 to 280) |
| Back problems | 14,642 | 12,080 | -17.5 (-19.5 to -15.5) | 5,676 | 5,952 | 4.9 (1.1 to 8.7) |
| Blood disorder | 823 | 504 | -38.8 (-45.2 to -31.6) | 463 | 399 | -13.8 (-24.6 to -1.5) |
| Cancer | 825 | 428 | -48.1 (-53.8 to -41.7) | 1,269 | 1,040 | -18.0 (-24.5 to -11.0) |
| Cardiac and circulatory | 3,315 | 2,811 | -15.2 (-19.4 to -10.8) | 1,924 | 2,117 | 10.0 (3.4 to 17.0) |
| Chest and respiratory | 12,175 | 8,034 | -34.0 (-35.8 to -32.1) | 3,831 | 7,347 | 91.8 (84.4 to 99.4) |
| Cough, flu | 69,177 | 30,507 | -55.9 (-56.5 to -55.3) | 4,071 | 9,966 | 145 (136 to 154) |
| Dental and oral problems | 6,489 | 4,862 | -25.1 (-27.8 to -22.2) | 562 | 458 | -18.5 (-28.0 to -7.8) |
| Ear, nose, throat | 16,346 | 9,840 | -39.8 (-41.3 to -38.3) | 2,909 | 2,391 | -17.8 (-22.1 to -13.2) |
| Endocrine, glandular problems | 1,212 | 814 | -32.8 (-38.5 to -26.6) | 618 | 502 | -18.8 (-27.8 to -8.6) |
| Eye problems | 4,673 | 2,998 | -35.8 (-38.7 to -32.8) | 1,313 | 785 | -40.2 (-45.3 to -34.7) |
| Gastrointestinal problems | 98,113 | 56,124 | -42.8 (-43.4 to -42.2) | 5,614 | 4,661 | -17.0 (-20.1 to -13.7) |
| Genitourinary, gynaecological | 11,977 | 9,199 | -23.2 (-25.3 to -21.1) | 4,175 | 2,545 | -39.0 (-42.0 to -36.0) |
| Headache, migraine | 33,292 | 33,648 | 1.1 (-0.5 to 2.6) | 1,309 | 1,763 | 34.7 (25.4 to 44.7) |
| Infectious diseases | 2,463 | 4,174 | 69.5 (61.2 to 78.1) | 897 | 3,681 | 310 (281 to 341) |
| Injury, fracture | 7,160 | 5,643 | -21.2 (-23.9 to -18.4) | 6,155 | 4,998 | -18.8 (-21.8 to -15.7) |
| Mental health | 14,973 | 15,768 | 5.3 (3.0 to 7.7) | 20,750 | 30,283 | 45.9 (43.4 to 48.5) |
| Nervous system disorders | 1,424 | 1,092 | -23.3 (-29.1 to -17) | 753 | 749 | -0.5 (-10.1 to 10.1) |
| Other musculoskeletal disorders | 18,215 | 13,741 | -24.6 (-26.2 to -22.9) | 10,204 | 8,004 | -21.6 (-23.8 to -19.2) |
| Pregnancy-related disorders | 7,110 | 5,618 | -21.0 (-23.7 to -18.2) | 2,800 | 3,640 | 30.0 (23.8 to 36.6) |
| Skin disorders | 3,201 | 2,762 | -13.7 (-18.0 to -9.2) | 1,054 | 1,196 | 13.5 (4.5 to 23.3) |
| Other and unknown | 50,694 | 28,155 | -44.5 (-45.3 to -43.6) | 11,711 | 9,579 | -18.2 (-20.4 to -16) |

<sup>a</sup>Calculated as 100\*(Number in 2020 – Number in 2019)/Number in 2019

Table 4 shows percentage changes from 2019 to 2020 in numbers of new absences during weeks 21-29, according to whether individuals had COVID-19 sickness absence during weeks 11-18 of 2020. When statistical uncertainty (as indicated by 95% confidence intervals) is taken into account, there were few clear indications of a differential change in subsequent patterns of new sickness absence following previous absence for COVID-19. However, the year-on-year increase in new absences for pregnancy-related disorders during weeks 21-29 was much greater among women with earlier COVID-19 sickness absence (215%, 95%CI 159% to 284%) than in those without COVID-19 (2.8%, 95%CI -1.8% to 7.6%).

**Table 4.** Changes from 2019 to 2020 in numbers of new episodes of sickness absence during weeks 21-29 by category of sickness absence, according to whether individuals had new COVID-19 sickness absence during weeks 11-18 of 2020.

| Category of sickness absence | No new COVID-19 sickness absence during Weeks 11-18 of 2020 |  |  | New COVID-19 sickness absence during Weeks 11-18 of 2020 |  |  |
| --- | --- | --- | --- | --- | --- | --- |
|  | Number in 2019 | Number in 2020 | <sup>a</sup>Percentage change (95%CI) | Number in 2019 | Number in 2020 | <sup>a</sup>Percentage change (95%CI) |
| All categories | 195,563 | 151,106 | -22.7 (-23.2 to -22.2) | 22,627 | 18,899 | -16.5 (-18.1 to -14.8) |
| All categories except COVID-19 | 195,563 | 140,839 | -28.0 (-28.5 to -27.5) | 22,627 | 16,690 | -26.2 (-27.7 to -24.7) |
| Asthma | 791 | 685 | -13.4 (-21.8 to -4.1) | 94 | 96 | 2.1 (-23.2 to 35.7) |
| Back problems | 8,798 | 8,605 | -2.2 (-5.1 to 0.8) | 1,061 | 1,017 | -4.1 (-12.0 to 4.5) |
| Blood disorder | 574 | 378 | -34.1 (-42.2 to -25) | 66 | 49 | -25.8 (-48.7 to 7.4) |
| Cancer | 990 | 620 | -37.4 (-43.4 to -30.8) | 63 | 58 | -7.9 (-35.6 to 31.5) |
| Cardiac and circulatory | 2,279 | 2,114 | -7.2 (-12.6 to -1.6) | 287 | 289 | 0.7 (-14.5 to 18.6) |
| Chest and respiratory | 6,183 | 2,836 | -54.1 (-56.1 to -52.0) | 680 | 462 | -32.1 (-39.6 to -23.5) |
| Cough, flu | 25,165 | 5,766 | -77.1 (-77.7 to -76.4) | 2,945 | 637 | -78.4 (-80.1 to -76.4) |
| Dental and oral problems | 3,059 | 2,600 | -15.0 (-19.3 to -10.4) | 359 | 329 | -8.4 (-21.1 to 6.4) |
| Ear, nose, throat | 8,111 | 4,858 | -40.1 (-42.2 to -37.9) | 963 | 519 | -46.1 (-51.6 to -40.0) |
| Endocrine, glandular problems | 799 | 564 | -29.4 (-36.6 to -21.4) | 88 | 90 | 2.3 (-23.8 to 37.2) |
| Eye problems | 2,578 | 1,814 | -29.6 (-33.7 to -25.3) | 300 | 187 | -37.7 (-48.1 to -25.2) |
| Gastrointestinal problems | 44,211 | 29,352 | -33.6 (-34.6 to -32.6) | 5,106 | 3,335 | -34.7 (-37.5 to -31.8) |
| Genitourinary, gynaecological | 7,103 | 5,656 | -20.4 (-23.1 to -17.5) | 861 | 671 | -22.1 (-29.5 to -13.8) |
| Headache, migraine | 15,617 | 17,038 | 9.1 (6.8 to 11.5) | 1,745 | 2,015 | 15.5 (8.3 to 23.1) |
| Infectious diseases | 1,505 | 1,313 | -12.8 (-19.0 to -6.1) | 185 | 141 | -23.8 (-38.8 to -5.1) |
| Injury, fracture | 5,922 | 5,212 | -12.0 (-15.2 to -8.7) | 712 | 597 | -16.2 (-24.8 to -6.5) |
| Mental health | 16,126 | 18,673 | 15.8 (13.4 to 18.3) | 1,848 | 2,119 | 14.7 (7.7 to 22.0) |
| Nervous system disorders | 974 | 843 | -13.4 (-21.1 to -5.1) | 108 | 107 | -0.9 (-24.2 to 29.4) |
| Other musculoskeletal disorders | 12,639 | 10,492 | -17.0 (-19.1 to -14.8) | 1,643 | 1,256 | -23.6 (-29.0 to -17.7) |
| Pregnancy-related disorders | 3,569 | 3,669 | 2.8 (-1.8 to 7.6) | 130 | 410 | 215 (159 to 28) |
| Skin disorders | 2,051 | 1,845 | -10.0 (-15.5 to -4.2) | 197 | 221 | 12.2 (-7.4 to 35.9) |
| Other and unknown | 26,519 | 15,906 | -40.0 (-41.2 to -38.8) | 3,186 | 2,085 | -34.6 (-38.1 to -30.8) |

## Discussion

Our analysis confirms that during the first wave of COVID-19 in England there were major changes in the incidence of sickness absence amongst NHS staff as compared with the corresponding period a year earlier. For some diagnostic categories (e.g., asthma, chest and respiratory disease, infectious diseases and mental illness) rates of absence increased (at least initially), whereas for others (e.g. musculoskeletal disorders, injury and fracture, gastrointestinal disease, genitourinary and gynaecological disease, and most notably, cancer), there were substantial reductions. COVID-19 sickness absence during weeks 11 to 18 of 2020, was not clearly associated with a higher year-on-year rise in new sickness absence during weeks 21 to 29, other than for pregnancy-related disorders.

### Methodological considerations

To our knowledge this is the first large study of the effects of the COVID-19 pandemic on sickness absence in healthcare workers for illness not directly attributable to coronavirus infection. The large sample size (almost a million individuals) gave the investigation high statistical power, and because we limited it to staff who were employed continuously throughout the study period, changes in numbers of new absences directly reflected changes in incidence rates. Moreover, they could not be confounded by differences between individuals in propensity to take sickness absence when ill, although they could reflect changes over time in thresholds for taking absence.

Information in the central ESR database had been collected prospectively through monthly updates, which were provided by NHS trusts in a standardised format. Dates of absence should have been highly reliable, but reasons for absence, which will have been determined originally from a combination of self-report and (for longer episodes) medical certification, may have been more prone to error, and also to inconsistencies in coding. In general, however, we would not expect there to have been systematic changes in the misclassification of reasons for sickness absence over the course of the study period.

The broad categories that were used when coding reasons for absence should have reduced the scope for misclassification, but they prevented us from exploring patterns of absence in finer detail. Nor was it possible to investigate longer term trends, although the year-on-year comparison for the weeks 2-10 provided some insight into levels of change that might have been expected in the absence of COVID-19.

The periods of the year that we studied were specified a priori, and such that public holidays (e.g. around Easter) fell in the same period in each year. The data were complete up to 31 July 2020, and by setting the end of the last period a little earlier in July, we ensured that we could reliably determine whether episodes had lasted for longer than 7 days.

### Interpretation of findings

Absences for infectious disease and chest and respiratory disorders increased sharply in March 2020 as compared with the previous year, and the rise closely paralleled the trajectory of COVID-19 sickness absence. This may have reflected failure to correctly identify and label some illness as COVID-related, especially early in the epidemic when testing was less widely available. In addition, individuals may have had a lower threshold for taking absence for illness with COVID-like symptoms because of the possibility that it might be caused by coronavirus. Year-on-year increases were predominantly for longer duration absences (Table 3), which suggests that the former was the main driver of the increase.

The pattern of sickness-absence attributed to cough/flu was different, with year-on-year reductions in sickness-absence episodes throughout the first wave of the epidemic. However, those reductions related only to short-duration episodes, and following the onset of the epidemic, new long-term episodes more than doubled (Table 3). The rise in longer duration absence for cough/flu is again likely to reflect failure to identify and label some illnesses as COVID-related. The fall in short-term absence may be attributable to reductions in the incidence of influenza and other common respiratory infections as a consequence of measures taken to reduce transmission of coronavirus. (11, 12)

The surge in new absences for asthma closely paralleled the rise in COVID-19 sickness absence (Figure 1) and was driven by episodes of longer duration (Table 3). It is possible that during the early phase of the epidemic, some workers with asthma took precautionary sickness absence because of concerns that they would be unusually vulnerable to COVID-19; at that stage the evidence on this issue was inconclusive.(13) A cross-sectional study of Danish healthcare workers found that fear of SARS-CoV-2 infection was common (prevalence ranging from 30% among those employed in psychiatry to 49% in ambulance workers)(14) and during pandemics, the willingness of healthcare workers to work can vary hugely according to the context within which they are expected to function, and perceptions of personal safety.(15)

The year-on-year fall in new episodes of absence for injury and fracture, which applied to both short and longer duration absence (Table 3), was more marked in the early phase of the epidemic (Table 2) when restrictions on activities outside work were greatest. It may have resulted, at least in part, from lower rates of injuries because of reductions in activities such as sports and driving. In reporting a fall in orthopaedic referrals and operative procedures for trauma during the COVID-19 epidemic, Murphy et al noted that the observed patterns of injury (low energy and fragility trauma persisted whilst injuries associated with younger people reduced) supported the view that it was in part a consequence of social distancing measures.(16)

A year-on-year increase in sickness absence for mental health (particularly that of longer duration) was apparent even before the COVID-19 epidemic took off, but it was most marked in weeks 11 to 20, suggesting that stresses relating to the epidemic (either at or outside work) may have led to an increase in mental illness. This will be explored in more detail in a separate paper.

New episodes of sickness absence for cardiovascular disease and cancer declined in the 20 weeks following the onset of the COVID-19 epidemic, but the reduction was much greater for cancer (30%) than for cardiac and circulatory disorders (6%). There is no reason to suspect that the incidence of such disease changed as a consequence of the pandemic, and the trends are more likely to have been driven by changes in health-seeking behaviour, and the postponement of less urgent investigations (e.g. in the follow-up of patients with previously diagnosed disease) due to reprioritisation of health systems.(17) There may have been some reluctance to seek medical advice about new symptoms when healthcare services were under pressure, especially where symptoms were not seriously incapacitating. In addition, individuals may have postponed medical consultation because they were preoccupied in adjusting to demands posed by the pandemic in their personal and professional lives. A contribution from changes in health-seeking behaviour would accord with the observation that the reduction in sickness absence for cancer was greatest for short-duration episodes. Whatever the reason, the finding adds to concerns about an impending problem from late diagnosis and treatment of cancers as a consequence of the pandemic. It has been estimated that referrals for suspected cancer in the UK during April to August of 2020 were down by 350,000 as compared with the same period in 2019,(18) and that as a consequence, 40,000 fewer patients commenced cancer treatment in 2020.(19) In the case of cardiovascular disease, the impact of delayed diagnosis and treatment may be more immediate than for cancer, and that could explain why the incidence of longer duration absence for cardiac and circulatory disorders rose despite a fall in short-duration absences (Table 3).

A combination of factors could have contributed to declines following the onset of the epidemic in absence for categories of illness such as dental and oral problems; disorders of ear, nose and throat; endocrine and glandular disease; eye problems; genitourinary and gynaecological disorders; diseases of the nervous system; and skin problems. They include: altered thresholds for taking absence for minor symptoms because of a wish to support patients and colleagues when services are stretched; diversion of resources from other services (e.g. less urgent surgery) to the management of COVID-19; and avoidance of healthcare settings because of a perceived risk of exposure to infection.(20) The last two could again be expected to forebode long-term challenges from a backlog of untreated morbidity.

Pregnancy-related disorders were the only category of sickness absence for which the year-on-year increase was greatest in weeks 21 to 29, and remarkably, that increase was limited to women who had an earlier episode of COVID-19 sickness absence during weeks 11 to 18 (Table 4). This cannot be explained by women opting for earlier maternity leave, which was coded separately from sickness absence. Nor is it likely to reflect generic fears about risks from COVID-19 during pregnancy. The rise occurred after the initial peak of the epidemic and did not follow the pattern observed for asthma. Moreover, it was limited to women with earlier COVID-19 sickness absence. The latter is an imperfect measure of true COVID-19 infection, but we have shown previously that it correlated with a positive antibody test for SARS-CoV-2.(9) Evidence is emerging that COVID-19 poses an increased risk in pregnancy, with higher odds of premature birth than in women who do not have the disease (21), and a greater risk of severe illness (particularly in the context of high body mass index and pre-existing comorbidities) as compared with that in infected women of the same age who are not pregnant.(22) However, it seems unlikely that such problems would account for an increase in absence for pregnancy-related disorders on the scale that we observed. To understand the phenomenon better, further research is needed to establish the exact circumstances and reasons for absence in a representative sample of cases.

## Conclusions

The COVID-19 pandemic has had a profound effect on patterns of sickness absence amongst NHS staff. The diverging trends that we report may have been driven by various mechanisms including: direct effects of COVID-19; a lower threshold for absence because of symptoms that might be caused by coronavirus infection; fears about vulnerability to COVID-19 in the presence of some comorbidities; pressures either at work or domestically as a consequence of the epidemic; a higher threshold for taking sickness absence in general because of the need to respond to the emergency posed by COVID-19; changes in activities outside work as a consequence of the epidemic; reluctance to present to medical services because of concerns about their being overloaded or the risk of exposure to infection; changes in the treatment of some disorders because of the diversion of resources to care of patients with COVID-19; and longer term trends unrelated to COVID-19.

Of particular concern is the marked reduction in sickness absence for cancer, which suggests an added burden of future morbidity, and perhaps mortality, as a consequence of delays in diagnosis and treatment. Such effects would be expected to extend to the wider population, and not to be confined only to NHS staff. In addition, plans are needed to manage a backlog of less urgent treatment for many other categories of disease that has been postponed because of the COVID-19 pandemic. The observed increase in sickness absence for mental illness will be explored in detail in a separate paper. Further research should be undertaken to understand the rise in absence for pregnancy-related disorders, which was limited to women with earlier COVID-19 sickness absence, possibly through detailed review of a representative case series.

## Data Availability

Data is available upon request from the NHS Employee Staff Record Warehouse at NHS England.

## Acknowledgments

We are very grateful to the following, without whom the study would not have been possible: Sam Wright, Workforce Information Advisor, NHS Electronic Staff Record, and Mike Vickerman, Workforce Information and Analysis, DHSC. Dr Gavin Debrera (Public Health England) and Dr Kit Harling gave invaluable help in planning the study.

We are grateful too, to Lee Isidore, Manal Sadik and Victoria Thorpe for their helpful input into the interpretation of our findings.

